# SARS-COV-2 antibody prevalence in patients on dialysis in the US in January 2021

**DOI:** 10.1101/2021.03.07.21252786

**Authors:** Shuchi Anand, Maria Montez-Rath, Jialin Han, LinaCel Cadden, Patti Hunsader, Russell Kerschmann, Paul Beyer, Scott D Boyd, Pablo Garcia, Mary Dittrich, Geoffrey A Block, Julie Parsonnet, Glenn M Chertow

## Abstract

**Background:** To estimate seroprevalence of SARS-CoV-2 antibodies in the US, the country with the world’s largest absolute numbers of COVID19 cases and deaths, we conducted a cross-sectional assessment from a sample of patients receiving dialysis in January 2021.

**Methods:** We tested remainder plasma of 21,424 patients receiving dialysis through the third-largest US dialysis organization, with facilities located nationwide. We used the Siemens spike protein receptor binding domain total antibody assay to estimate crude SARS-CoV-2 seroprevalence, and then estimated seroprevalence for the US dialysis and adult population by standardizing by age, sex and region. We also compared January 2021 seroprevalence and case-detection rates to that from a similar subsample of patients receiving dialysis who had been tested in July 2020.

**Results:** Patients in the sample were disproportionately from older age and minority race/ethnic groups. Seroprevalence of SARS-CoV-2 was 18.9% (95% CI: 18.3-19.5%) in the sample, 18.7% (18.1-19.2%) standardized to the US dialysis population, and 21.3% (20.3-22.3%) standardized to the US adult population (range 15.3-20.8% in the Northeast and South respectively). Younger age groups (18-44 years), and persons self-identifying as Hispanic or living in Hispanic neighborhoods, and persons living in the poorest neighborhoods were among the subgroups with the highest seroprevalence (25.9% (24.1-27.8%), 25.1% (23.6-26.4%), 24.8% (23.2-26.5%) respectively). Compared to data from July 2020, we observed diminished variability in seroprevalence by geographic region and urban-rural status. Estimated case detection rate increased from 14% to 23% in July 2020 to January 2021.

**Conclusions:** A year after the first case of SARS-CoV-2 infection was detected in the US, fewer than one in four adults have evidence of SARS-CoV-2 antibodies. Vaccine roll out to majority minority neighborhoods and poorer neighborhoods will be critical to disrupting the spread of infection.

**Funding:** Ascend Clinical Laboratories funded remainder-plasma testing.

## Introduction

In the year-long period since SARS-CoV-2 case was reported in the US, COVID-19 cases, deaths, and hospitalizations overwhelmed health systems in the country. To date, more than 25 million cases and 500,000 deaths have been attributed to SARS-CoV-2 infection^1,2^. While in Spring 2020, the most intense burden of disease was experienced in and around the state of New York, nearly all regions in the US including rural areas have recently faced threats to their hospital capacity, with the largest numbers accruing between November and December 2020^2^.

Data continue to support that symptomatic cases represent a fraction of persons infected with SARS-CoV-2^3^. The vast majority of persons with infection, however, whether asymptomatic or symptomatic, do mount a specific antibody response^4-6^, with the latest data indicating SARS-CoV-2 receptor binding domain (RBD) IgG antibodies persist for at least four to six months after infection^4,7,8^. Thus, seroprevalence estimates remain an essential measure of the extent of SARS-CoV-2 community spread.

Following our proposed strategy of performing repeated seroprevalence surveys among patients on dialysis, a population broadly representative of groups susceptible to SARS-CoV-2 infection and among whom routine requirements for monthly laboratories facilitates surveillance^9^, we implemented a cross-sectional seroprevalence analysis in a sample of 21,424 patients on dialysis in the US in January 2021. Our goal was to assess SARS-CoV-2 seroprevalence prior to widespread roll out of COVID19 vaccination. We also tested the persistence of previously observed differences in seroprevalence by region, age, sex, race/ethnicity, and other community-level strata of interest including population density and neighborhood poverty. Finally, to assess for improvements in diagnosis and treatment since our July survey, we compared case detection attributed to COVID19 to our seroprevalence estimates at the July 2020 and January 2021 timepoints.

## Methods

We replicated methods presented previously^9^, including the use of a highly sensitive and specific total RBD total antibody assay on remnant plasma from routine monthly laboratory draws. The primary difference was in the sample population. Previously, samples were obtained from multiple dialysis networks and from patients residing in 46 states. The current sample derived from a single national dialysis network, US Renal Care, the third largest dialysis center in the US with 518 facilities and from patients residing in 43 states. We tested all patients (n=21424) within this single dialysis network in preparation for a longitudinal study being conducted in this network to ascertain response to COVID19 vaccination.

The study received Institutional Review Board Approval from Stanford University.

### Assay characteristics

In partnership with a commercial laboratory receiving routine monthly laboratories of patients on dialysis, we tested the remnant plasma of all patients receiving dialysis through US Renal Care. We used the Siemens RBD total antibody (Ig) chemiluminescence assay, which has 100% sensitivity (≥14 days post +PCR test) and 99.8% specificity^10^.

### Statistical analyses

We determined seroprevalence estimates and 95% confidence intervals for the crude (unweighted) sample from January 2021. We then standardized seroprevalence estimates accounting for age, sex, and region, using the distribution of the US dialysis population as available through the United States Renal Data System^11^ and for the US adult population as available through the 2018 American Community Survey 1-year estimates^12^. Next we examined differences in seroprevalence by the following strata: age, sex, region, self-reported race/ethnicity, neighborhood race/ethnicity composition, neighborhood poverty, and urban/rural status^9^.

We also compared data on differences in seroprevalence by patient and neighborhood characteristics and region of residence strata to the observed differences in July 2020. To ensure accurate comparisons, we limited the July 2020 sample to those obtained within US Renal Care (n=11,746 (41%) of 28,503 tested in July,**supplemental Table 1**).

Finally, we compared case detection and infection fatality rates in July 2020 versus January 2021. We computed case detection rates as the proportion of detected cases per 100,000^2^ from a denominator totaling seroprevalence estimates per 100,000 (standardized to the US adult population) and deaths per 100,000 in January 1 2020-June 31 2020, and July 1 2020-December 31 2020 for the July 2020 and January 2021 estimates respectively. We adjusted the numerator of detected cases to reflect adults only using Centers for Disease Control data on proportion of overall cases occurring in adults (∼92% in June 2020^13^ and ∼89% in December 2020^14^). We computed the infection fatality rate as the proportion of deaths per 100,000 from a denominator of seroprevalent persons per 100,000 in the corresponding time periods.

## Results

Of the 21,424 persons tested in the first week of January 2021, a majority were men and were older than 65 years of age, similar to the overall US dialysis population (**Table 1**). Although patients resided in a total of 43 states, 33 states contributed >30 patients to the sample. Persons from the South and West regions were modestly overrepresented. Neighborhood race/ethnicity composition closely matched the US dialysis population. Compared to the US adult population, sampled patients were older, more likely male, and more likely to be of non-white race and Hispanic ethnicity.

**Table 1.** Comparison of sampled population, US adult dialysis population, and US adult population (January 2021)

| Patient characteristics | Sample population<br>(January 2021)<br>N=21,424 |  | US adult<br>dialysis<br>population<br>N=499,150 |  | US adult<br>population<br>N=253,815,197 |  |
| --- | --- | --- | --- | --- | --- | --- |
|  | Count | % | Count | % | Count | % |
| **Age** |  |  |  |  |  |  |
| 18-44 | 2314 | 11 | 60,540 | 12 | 117,499,477 | 46 |
| 45-64 | 8635 | 40 | 207,022 | 41 | 83,892,606 | 33 |
| 65-79 | 7847 | 37 | 174,341 | 35 | 39,949,825 | 16 |
| ≥80 | 2668 | 12 | 57,247 | 11 | 12,473,289 | 5 |
| **Sex** |  |  |  |  |  |  |
| M | 12,265 | 57 | 285,281 | 57 | 123,578,869 | 49 |
| F | 9199 | 43 | 213,869 | 43 | 130,236,328 | 51 |
| **Race and Ethnicity\*<sup>&</sup>** |  | <sup>&</sup> |  |  |  |  |
| Hispanic | 2945 | 18 | 87,611 | 18 | 60,861,275 | 19 |
| Non-Hispanic white | 6311 | 37 | 203,421 | 41 | 197,202,727 | 60 |
| Non-Hispanic Black | 4875 | 29 | 173,190 | 35 | 39,717,152 | 12 |
| Non-Hispanic Other | 2834 | 17 | 34,928 | 7 | 28,493,202 | 9 |
| **ZCTA Majority Race and Ethnicity\*<sup>^+</sup>** |  |  |  |  |  |  |
| Non-Hispanic white | 6481 | 30 | 206,678 | 41 | 189,968,192 | 58 |
| Non-Hispanic Black | 1504 | 7 | 54,999 | 11 | 12,550,083 | 4 |
| Hispanic | 4023 | 19 | 52,953 | 11 | 26,310,796 | 8 |
| Hispanic and Black | 2321 | 11 | 43,396 | 9 | 17,238,911 | 5 |
| Integrated | 7135 | 33 | 140,781 | 28 | 80,206,374 | 25 |
| **Region** |  |  |  |  |  |  |
| Northeast | 2113 | 10 | 78,619 | 16 | 44,519,465 | 18 |
| South | 10857 | 50 | 214,974 | 43 | 96,250,597 | 38 |
| Midwest | 2055 | 10 | 94,490 | 19 | 52,876,708 | 21 |
| West | 6439 | 30 | 111,067 | 22 | 60,168,427 | 24 |

US adult population in 2018, US adult patients on dialysis population as of January 1 2017.
\*Race, ethnicity, and ZCTA Majority Race and Ethnicity computed for total U.S. 2018 population N=326,274,356.
<sup>&</sup>Proportions are reported for persons with available data; 4505 (21) in the sample were missing race and ethnicity.
<sup>^</sup>343 and 583 people in the U.S. Renal Data System and sample populations missing data on ZCTA Majority Race/Ethnicity due to missing zip code respectively. <sup>+</sup>ZCTA Majority defined as population in ZCTA ≥ 60 Hispanic, Non-Hispanic Black, or Non-Hispanic White; if in remainder ZCTAs Hispanic and Black population exceeded ≥60, ZCTA defined as 'Hispanic and Black', else as 'Other'.
Abbreviations: ZCTA-zip code tabulation area

In January 2021, SARS-CoV-2 RBD seroprevalence estimates were 18.9% (95% CI 18.3, 19.5%) in the sample, 18.7% (18.1, 19.2%) standardized to US dialysis, and 21.3% (20.3, 22.3%) standardized to US adult population respectively (**Table 2**). Younger age groups (18-44 years), and persons self-identifying as Hispanic or living in Hispanic neighborhoods, and persons living in the poorest neighborhoods were among the subgroups with the highest seroprevalence.

**Table 2.**
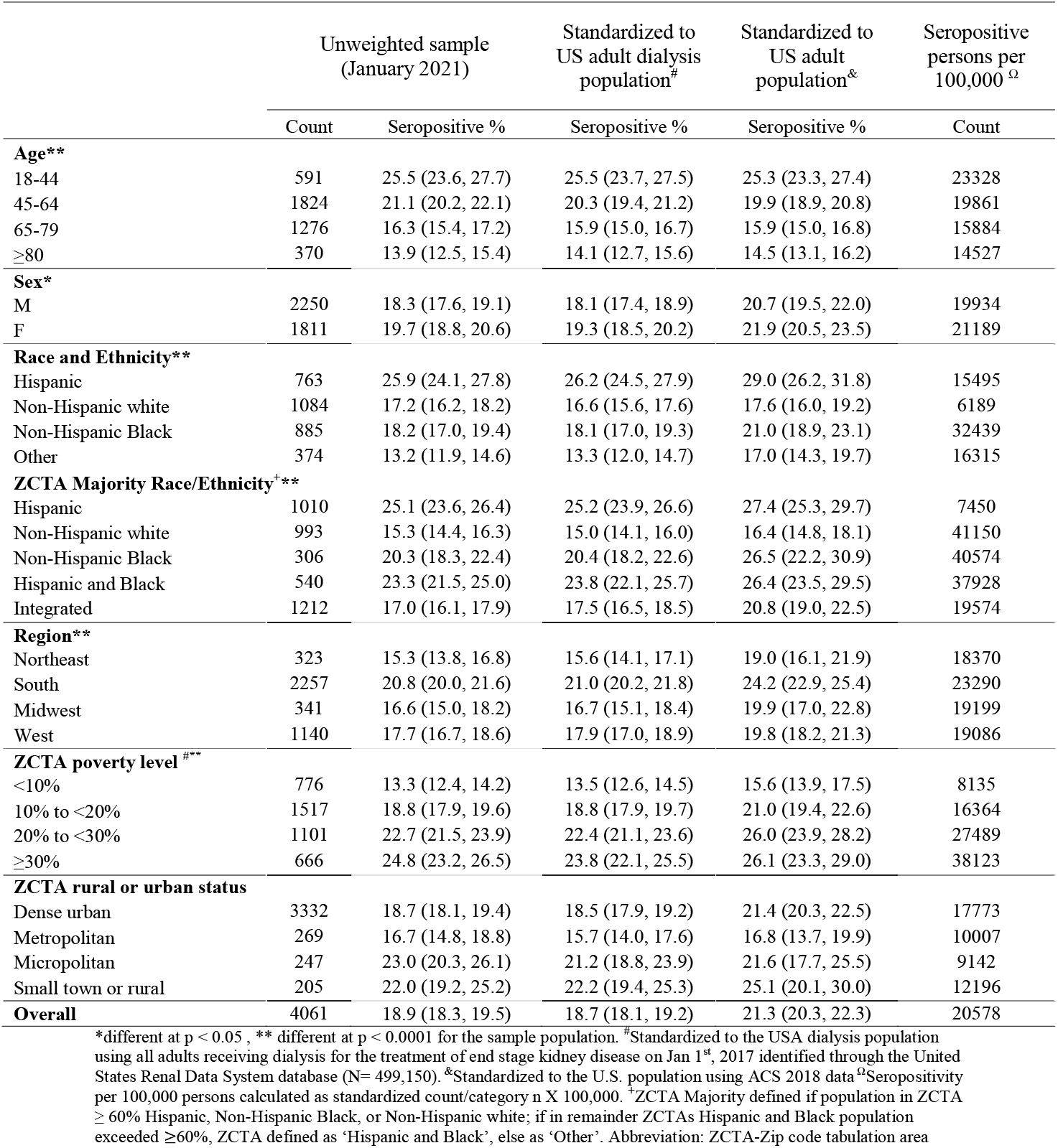
Seroprevalence of SARS-CoV2 antibodies in the study sample and standardized to US dialysis and US adult population in January 2021

|  | Unweighted sample<br>(January 2021) |  | Standardized to<br>US adult dialysis<br>population<sup>#</sup> | Standardized to<br>US adult<br>population<sup>&</sup> | Seropositive<br>persons per<br>100,000<sup>Ω</sup> |
| --- | --- | --- | --- | --- | --- |
|  | Count | Seropositive % | Seropositive % | Seropositive % | Count |
| **Age\*\*** |  |  |  |  |  |
| 18-44 | 591 | 25.5 (23.6, 27.7) | 25.5 (23.7, 27.5) | 25.3 (23.3, 27.4) | 23328 |
| 45-64 | 1824 | 21.1 (20.2, 22.1) | 20.3 (19.4, 21.2) | 19.9 (18.9, 20.8) | 19861 |
| 65-79 | 1276 | 16.3 (15.4, 17.2) | 15.9 (15.0, 16.7) | 15.9 (15.0, 16.8) | 15884 |
| ≥80 | 370 | 13.9 (12.5, 15.4) | 14.1 (12.7, 15.6) | 14.5 (13.1, 16.2) | 14527 |
| **Sex\*** |  |  |  |  |  |
| M | 2250 | 18.3 (17.6, 19.1) | 18.1 (17.4, 18.9) | 20.7 (19.5, 22.0) | 19934 |
| F | 1811 | 19.7 (18.8, 20.6) | 19.3 (18.5, 20.2) | 21.9 (20.5, 23.5) | 21189 |
| **Race and Ethnicity\*\*** |  |  |  |  |  |
| Hispanic | 763 | 25.9 (24.1, 27.8) | 26.2 (24.5, 27.9) | 29.0 (26.2, 31.8) | 15495 |
| Non-Hispanic white | 1084 | 17.2 (16.2, 18.2) | 16.6 (15.6, 17.6) | 17.6 (16.0, 19.2) | 6189 |
| Non-Hispanic Black | 885 | 18.2 (17.0, 19.4) | 18.1 (17.0, 19.3) | 21.0 (18.9, 23.1) | 32439 |
| Other | 374 | 13.2 (11.9, 14.6) | 13.3 (12.0, 14.7) | 17.0 (14.3, 19.7) | 16315 |
| **ZCTA Majority Race/Ethnicity<sup>+</sup>\*\*** |  |  |  |  |  |
| Hispanic | 1010 | 25.1 (23.6, 26.4) | 25.2 (23.9, 26.6) | 27.4 (25.3, 29.7) | 7450 |
| Non-Hispanic white | 993 | 15.3 (14.4, 16.3) | 15.0 (14.1, 16.0) | 16.4 (14.8, 18.1) | 41150 |
| Non-Hispanic Black | 306 | 20.3 (18.3, 22.4) | 20.4 (18.2, 22.6) | 26.5 (22.2, 30.9) | 40574 |
| Hispanic and Black | 540 | 23.3 (21.5, 25.0) | 23.8 (22.1, 25.7) | 26.4 (23.5, 29.5) | 37928 |
| Integrated | 1212 | 17.0 (16.1, 17.9) | 17.5 (16.5, 18.5) | 20.8 (19.0, 22.5) | 19574 |
| **Region\*\*** |  |  |  |  |  |
| Northeast | 323 | 15.3 (13.8, 16.8) | 15.6 (14.1, 17.1) | 19.0 (16.1, 21.9) | 18370 |
| South | 2257 | 20.8 (20.0, 21.6) | 21.0 (20.2, 21.8) | 24.2 (22.9, 25.4) | 23290 |
| Midwest | 341 | 16.6 (15.0, 18.2) | 16.7 (15.1, 18.4) | 19.9 (17.0, 22.8) | 19199 |
| West | 1140 | 17.7 (16.7, 18.6) | 17.9 (17.0, 18.9) | 19.8 (18.2, 21.3) | 19086 |
| **ZCTA poverty level<sup>\*\*\*</sup>** |  |  |  |  |  |
| <10% | 776 | 13.3 (12.4, 14.2) | 13.5 (12.6, 14.5) | 15.6 (13.9, 17.5) | 8135 |
| 10% to <20% | 1517 | 18.8 (17.9, 19.6) | 18.8 (17.9, 19.7) | 21.0 (19.4, 22.6) | 16364 |
| 20% to <30% | 1101 | 22.7 (21.5, 23.9) | 22.4 (21.1, 23.6) | 26.0 (23.9, 28.2) | 27489 |
| ≥30% | 666 | 24.8 (23.2, 26.5) | 23.8 (22.1, 25.5) | 26.1 (23.3, 29.0) | 38123 |
| **ZCTA rural or urban status** |  |  |  |  |  |
| Dense urban | 3332 | 18.7 (18.1, 19.4) | 18.5 (17.9, 19.2) | 21.4 (20.3, 22.5) | 17773 |
| Metropolitan | 269 | 16.7 (14.8, 18.8) | 15.7 (14.0, 17.6) | 16.8 (13.7, 19.9) | 10007 |
| Micropolitan | 247 | 23.0 (20.3, 26.1) | 21.2 (18.8, 23.9) | 21.6 (17.7, 25.5) | 9142 |
| Small town or rural | 205 | 22.0 (19.2, 25.2) | 22.2 (19.4, 25.3) | 25.1 (20.1, 30.0) | 12196 |
| **Overall** | 4061 | 18.9 (18.3, 19.5) | 18.7 (18.1, 19.2) | 21.3 (20.3, 22.3) | 20578 |

\*different at $p < 0.05$ , \*\* different at $p < 0.0001$ for the sample population. <sup>#</sup>Standardized to the USA dialysis population using all adults receiving dialysis for the treatment of end stage kidney disease on Jan 1<sup>st</sup>, 2017 identified through the United States Renal Data System database (N= 499,150). <sup>&</sup>Standardized to the U.S. population using ACS 2018 data<sup>Ω</sup> Seropositivity per 100,000 persons calculated as standardized count/category $n \times 100,000$ . <sup>+</sup>ZCTA Majority defined if population in ZCTA ≥ 60% Hispanic, Non-Hispanic Black, or Non-Hispanic white; if in remainder ZCTAs Hispanic and Black population exceeded ≥60%, ZCTA defined as 'Hispanic and Black', else as 'Other'. Abbreviation: ZCTA-Zip code tabulation area

We compared these values to observations from patients within the US Renal Care network tested in July 2020. In this subset in July, the sample, dialysis adjusted and US population adjusted seroprevalence rates were 4.4 (4.0, 4.8), 4.7 (4.3, 5.2), and 5.4 (4.6, 6.2), respectively (**supplemental Table 2**). Relative to July, January 2021 seroprevalences were 2-fold, 4-fold, and 5-fold higher in the Northeast, Midwest, and South and West respectively (**Figure 1**). Regional and rural vs. urban seroprevalences varied less than previously (**supplemental Figure 1**), but striking differences remained by race/ethnicity and poverty. Seroprevalence was 1·7-fold (95% CI 1·6, 1·9) higher among persons living in majority-Hispanic compared to majority-white neighborhoods, and 2·0-fold (95% CI 1·8, 2·3) higher among persons living in neighborhoods with ≥ 30% versus <10% of residents living in poverty (**Figure 2**).

**Figure 1:**
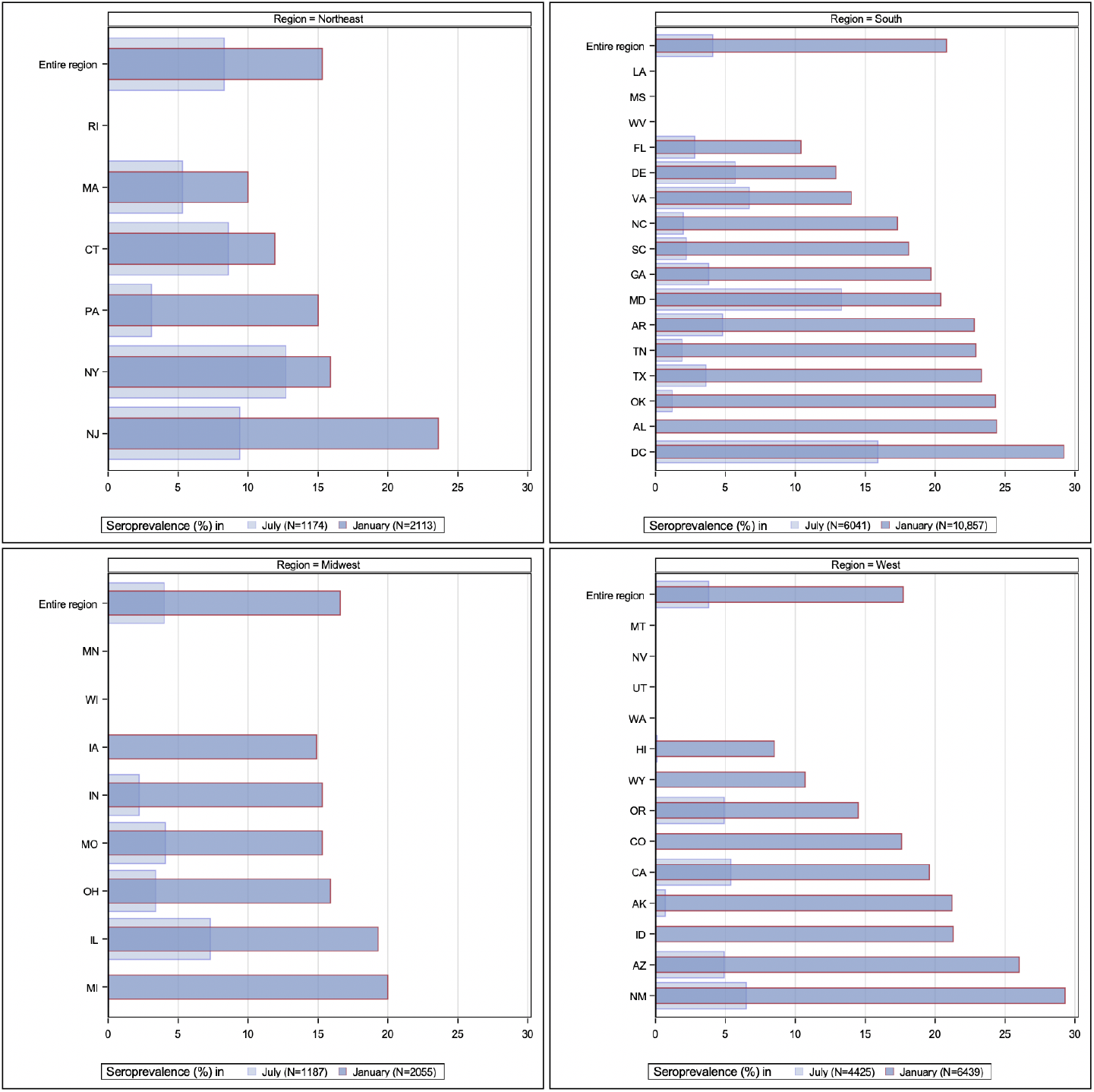
SARS-CoV-2 seroprevalence by US region in January 2021, with comparison to sampled states between July 2020 Overall seroprevalence was between 15-21% across all regions, with the highest seroprevalence in the US South. Compared to July data from patients dialyzing in the same dialysis facilities, seroprevalence was 1.8-fold (95% CI 1.5, 2.3) higher in the Northeast, 4.1-fold (95% CI 3.0, 5.7) higher in the Midwest and 4.6-fold (95% CI 3.9, 5.5) higher in the West. The largest increase in seroprevalence was observed in the South (5.1-fold (95% CI 4.5, 5.8)). All 43 states in which patients resided are listed in the figure but data are presented only for states with at least 30 persons in the January sample (n=33).

**Figure 2:**
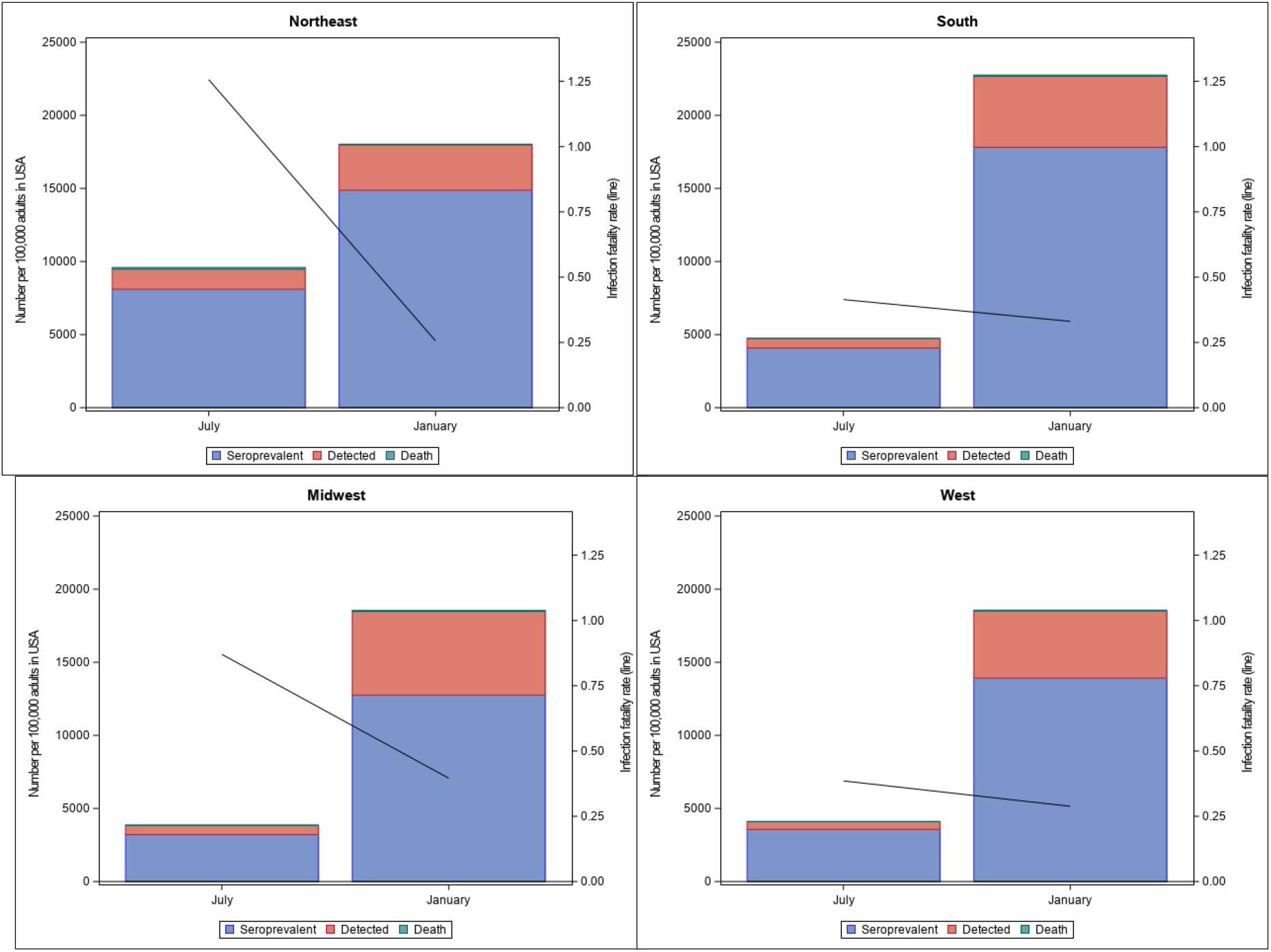
Case detection and infection fatality rates in January 2021, with comparison to July 2020 The detected cases per 100,000 between June-December 2020 were 23% of estimated seroprevalent persons and deaths attributed to COVID19 in January 2021 (range by region 17-30% in the Northeast and Midwest respectively). This proportion was an increase from detected cases being 14% of estimated seroprevalent persons and deaths attributed to COVID19 in July 2020 (range by region 13% to 16% in the West and Midwest respectively). The infection fatality rate in January 2021 was 0.3% (range 0.3-0.4%), compared with 0.7% in July 2020 (range 0.4-1.3%).

## Discussion

In a January, 2021 nationwide surveillance of SARS-CoV-2 seroprevalence among patients on dialysis, we found that, despite national surges in cases and almost half a million deaths, fewer than one in four US adults had evidence of SARS-CoV-2 antibody. Compared to July 2020, spread of SARS-CoV-2 throughout the US became more uniform across geographic regions, and urban and rural areas. The largest differences in seroprevalences occurred between younger and older age groups, residents of majority Hispanic versus majority white neighborhoods, and residents of poorer versus more affluent neighborhoods. Case detection improved but close to three in four cases remained undiagnosed. Our data emphasize the critical need for prompt, wide-spread vaccination, and ongoing mitigation efforts since on the basis of this seroprevalence evaluation, much of the population still remains vulnerable to SARS-CoV-2 infection.

Few up-to-date data exist on SARS-CoV-2 seroprevalence from countries with high numbers of cases and deaths from COVID19. In the US, the Centers for Disease Control (CDC) implemented a large and geographically diverse longitudinal sero-surveillance using repeated commercial laboratory testing. Our July seroprevalence estimates were in line with those reported using the CDC strategy.^15^ The latest available data from the CDC, however, are from November 2020, prior to the largest surge of cases in the US. Outside of the US, India performed over 15000 home visits in 700 communities testing 29,000 people with plasma nucleocapsid IgG testing between August and September 2020 and estimated seroprevalence of 7% among adults, at a time when the country had recorded roughly six million cases^16^. In the UK, the Real-time assessment of Community Transmission (REACT) study mailed lateral flow immunoassays to randomly selected UK residents^17,18^. This ongoing study has a wide reach (over 350,000 participants reached by fall 2020), amenable to repeat serosurveillance. In the last iteration prior to vaccine roll out, the authors observed a decline in seroprevalence over time possibly due to rapid decay in the qualitative measure of anti-Spike antibody, limiting the study’s ability to estimate overall infection rates in the UK^17^. Brazilian investigators using blood donor samples in Manaus, Brazil, created a seroprevalence model that incorporated a parameter for antibody decay. Using this strategy, they concluded that 75% in Manaus had been infected by October 2020.^19^ A later surge of infections in Manaus, however, has raised questions about model assumptions^20^.

Ease of replication, unbiased sampling, and high-quality immunoassays—all features of national dialysis sampling-are crucial to rapid and informative seroprevalence. Nonetheless, our strategy may also have modestly underestimated SARS-CoV-2 cumulative infections for two reasons. First, SARS-CoV-2 antibodies may wane over time. Several large studies, however, including our own performed in this population, indicate that the RBD domain antibody typically persists for at least 4-6 months^4,5^, longer than the nucleocapsid assays used in other seroprevalence studies^16,19^. Second, our current sample underrepresents New York City, a high-risk area, perhaps contributing to a lower seroprevalence than might be otherwise expected in the Northeast region.

Our data reliably assess differences in seroprevalence by patient and community-level characteristics over time. We note the rapid spread of the infection throughout the breadth of the US. Whereas in July 2020 seroprevalence rates varied from <1 to 34%, in January 2021, nearly all sampled states had seroprevalence estimates exceeding 10% and among the four regions, the range of seroprevalence was narrow (15-21%). These trends fit with other descriptions of the pandemic, for example in Brazil, where international travel first brought the pandemic to major cities with rapid spread in densely populated areas, but eventual spread occurred throughout the country^21^.

Our estimates for case detection and infection fatality—with infection fatality exceeding 1% in the Northeast after the first wave in the Spring—match modeled estimates by Yang et al.^22^ The improvement in both parameters since then indicate better detection and treatment, and are generally uniformly spread throughout the US.

Strengths of our study include its unbiased sampling of hard-to-reach populations, reflective of persons most vulnerable to SARS-CoV-2, and the use of a highly specific and sensitive assay for an antibody for which data indicate a prolonged persistence in a majority of infected persons^4,5,23^. This pragmatic approach enables rapid and repeated evaluations. The limitations include lack of data on SARS-CoV-2 rtPCR testing or COVID19 symptoms, and relative under-sampling from the Northeast region.

In summary, using unbiased data from a sentinel population, we estimate that fewer than one in four persons in the US had evidence of SARS-CoV-2 antibody in January 2021, well below the level needed to confer herd immunity. Residents of majority minority and poorer neighborhoods, and younger age groups have substantially higher prevalence of SARS-CoV-2 antibodies. Since these sub-populations overlap with persons expressing high levels of vaccine hesitancy in the US^24,25^, vaccination campaigns will need to engage these high-risk groups in order to achieve sufficient penetration to reach community-level protection against SARS-CoV-2.

## Data Availability

The dataset is expected to be available upon review of request.

## Author Contributions

SA assisted with cleaning and analysis planning, and manuscript writing. MMR developed analysis plan, supervised data analysis and contributed to manuscript writing. JH and PG undertook data cleaning and analysis, including linkage to external data and figure generation, and contributed to manuscript writing. LC and PH undertook sample processing and data preparation. GB and MD participated in study planning and sample and de-identified data provision. RK selected seroprevalence testing, supervised sample processing, and contributed to manuscript writing. PB co-conceived study, and secured seroprevalence testing. JP supervised study analysis plan, identified relevant external data, data interpretation, and supervised manuscript writing. SB assisted with data interpretation. GMC co-conceived study, supervised study analysis plan, and co-wrote manuscript.

---

**Supplemental Figure 1:**
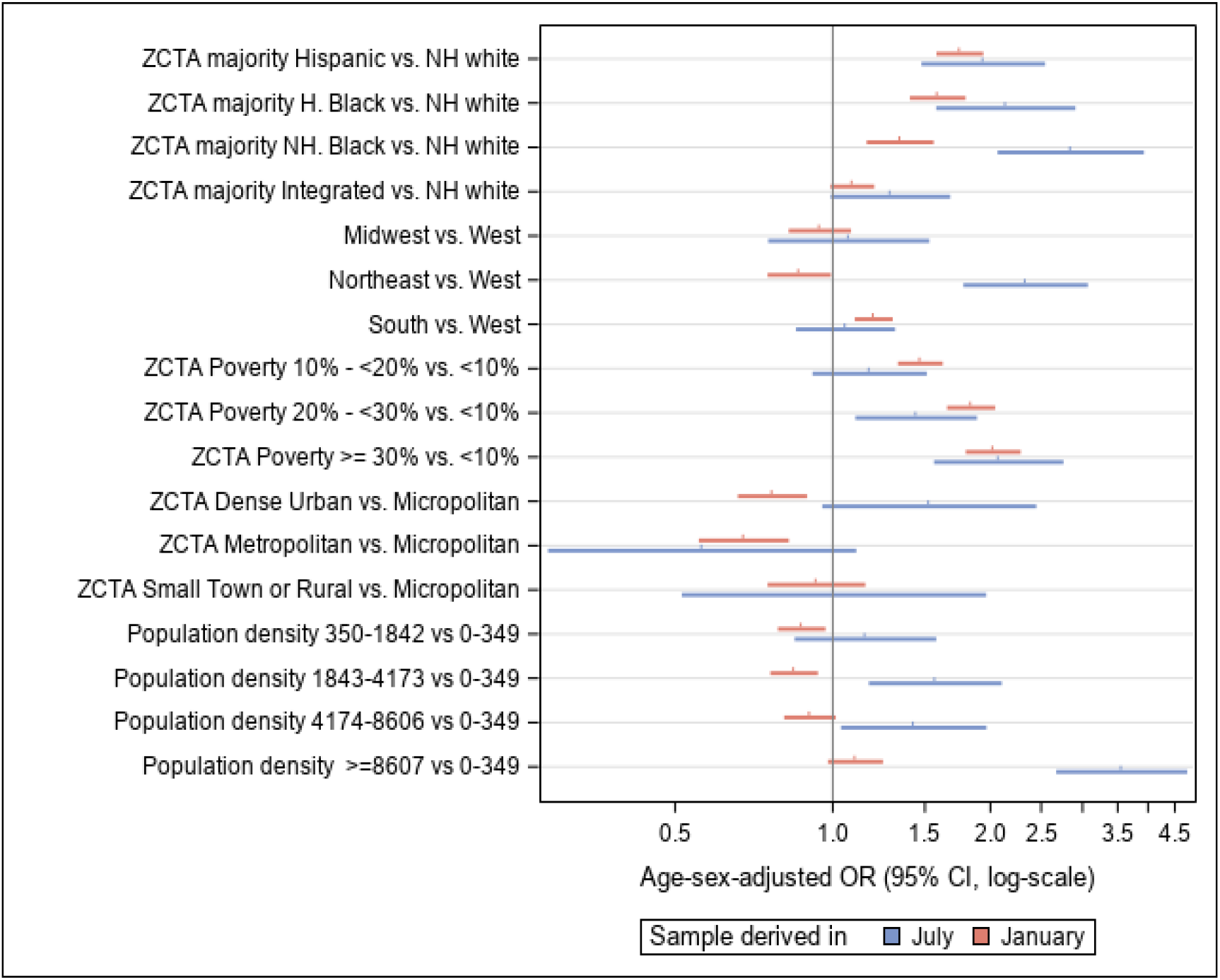
Neighborhood level characteristics associated with SARS-CoV-2 seroprevalence in January 2021, with comparisons to July 2021.

