## Supplemental Tables1&2, Figure1 for "SARS-COV-2 antibody prevalence in patients on dialysis in the US in January 2021"


**Supplemental Table 1** Comparison of sampled population, US adult dialysis population, and US adult population (July US Renal sample)
**p2**

**Supplemental Table 2** Seroprevalence of SARS-CoV2 antibodies in patients receiving dialysis in the US (July US Renal population)
**p3**

**Supplemental Figure 1** Differences in seroprevalence by patient and neighborhood characteristics between July 2020 and January 2021
**p4**

**Supplemental Table 1** Comparison of sampled population, US adult dialysis population, and US adult population (July US Renal sample)

| Patient characteristics | US Renal population July 2020 N=11 746 | | U.S. adult dialysis population N=499 150 | | U.S. adult population N=253 815 197 | |
|---|---|---|---|---|---|---|
| | Count | % | Count | % | Count | % |
| **Age** | | | | | | |
| 18-44 | 1304 | 11 | 60 540 | 12 | 117 499 477 | 46 |
| 45-64 | 4723 | 40 | 207 022 | 41 | 83 892 606 | 33 |
| 65-79 | 4298 | 37 | 174 341 | 35 | 39 949 825 | 16 |
| ≥80 | 1421 | 12 | 57 247 | 11 | 12 473 289 | 5 |
| **Sex** | | | | | | |
| Men | 6634 | 56 | 285 281 | 57 | 123 578 869 | 49 |
| Women | 5112 | 44 | 213 869 | 43 | 130 236 328 | 51 |
| **Race and Ethnicity*&** | | | | | | |
| Hispanic | 1902 | 18 | 87 611 | 18 | 60 861 275 | 19 |
| Non-Hispanic white | 4226 | 39 | 203 421 | 41 | 197 202 727 | 60 |
| Non-Hispanic Black | 3042 | 28 | 173 190 | 35 | 39 717 152 | 12 |
| Non-Hispanic Other | 1672 | 15 | 34 928 | 7 | 28 493 202 | 9 |
| **ZCTA Majority Race and Ethnicity*^+** | | | | | | |
| Non-Hispanic white | 3538 | 30 | 206 678 | 41 | 189 968 192 | 58 |
| Non-Hispanic Black | 848 | 7 | 54 999 | 11 | 12 550 083 | 4 |
| Hispanic | 2250 | 19 | 52 953 | 11 | 26 310 796 | 8 |
| Hispanic and Black | 1282 | 11 | 43 396 | 9 | 17 238 911 | 5 |
| Integrated | 3828 | 33 | 140 781 | 28 | 80 206 374 | 25 |
| **Region** | | | | | | |
| Northeast | 1172 | 10 | 78 619 | 16 | 44 519 465 | 18 |
| South | 5926 | 51 | 214 974 | 43 | 96 250 597 | 38 |
| Midwest | 1072 | 9 | 94 490 | 19 | 52 876 708 | 21 |
| West | 3576 | 30 | 111 067 | 22 | 60 168 427 | 24 |

U.S. adult population in 2018, U.S. adult patients on dialysis population as of January 1 2017.
*Race, ethnicity, and ZCTA Majority Race and Ethnicity computed for total U.S. 2018 population N=326,274,356. &Proportions are reported for persons with available data; 904 (7.7%) in the sample were missing race and ethnicity.
^343 and 583 people in the U.S. Renal Data System and sample populations missing data on ZCTA Majority Race/Ethnicity due to missing zip code respectively. +ZCTA Majority defined as population in ZCTA ≥ 60% Hispanic, Non-Hispanic Black, or Non Hispanic White; if in remainder ZCTAs Hispanic and Black population exceeded ≥60%, ZCTA defined as ‘Hispanic and Black’, else as ‘Other’.
Abbreviations: ZCTA-zip code tabulation area

**Supplemental Table 2** Seroprevalence of SARS-CoV-2 antibodies in patients receiving dialysis in the US (July US Renal population)

| | Unweighted sample (US Renal population July 2020) | | Standardized to US adult dialysis population# | Standardized to US adult population& | Seropositive persons per 100,000 Ω |
|---|---|---|---|---|---|
| | Count | Seropositive % | Seropositive % | Seropositive % | |
| **Age**** | | | | | |
| 18-44 | 74 | 5·7 (4·4, 6·9) | 6·1 (4·5, 7·7) | 6·1 (4·5, 7·7) | 5857 |
| 45-64 | 231 | 4·9 (4·3, 5·5) | 5·3 (4·5, 6) | 5·3 (4·5, 6) | 5230 |
| 65-79 | 163 | 3·8 (3·2, 4·4) | 4·2 (3·5, 4·9) | 4·2 (3·5, 4·9) | 4162 |
| ≥80 | 51 | 3·6 (2·6, 4·6) | 4·1 (2·8 5·3) | 4·1 (2·8 5·3) | 3984 |
| **Sex*** | | | | | |
| M | 285 | 4·3 (3·8, 4·8) | 5·3 (4.3, 6·3) | 5·3 (4.3, 6·3) | 5025 |
| F | 234 | 4·6 (4, 5·2) | 5·5 (4·4, 6·7) | 5·5 (4·4, 6·7) | 5544 |
| **Race and Ethnicity**** | | | | | |
| Hispanic | 120 | 6·3 (5·2, 7·4) | 6·9 (5·7, 8·2) | 8·0 (5·8, 10·1) | 5050 |
| Non-Hispanic white | 116 | 2·7 (2·3, 3·2) | 3 (2·4, 3·5) | 3·7 (2·6, 4·8) | 1613 |
| Non-Hispanic Black | 184 | 6 (5·2, 6·9) | 6·4 (5·5, 7·4) | 6·7 (5, 8·4) | 12367 |
| Other | 34 | 2 (1·4, 2·7) | 2·6 (1·7, 3·6) | 2·4 (1·2, 3·5) | 2492 |
| **ZCTA Majority Race and Ethnicity**+** | | | | | |
| Hispanic | 126 | 5·6 (4·7, 6·6) | 6.4 (5.2, 7.5) | 8·0 (5·9, 10·0) | 12308 |
| Non-Hispanic white | 102 | 2·9 (2·3, 3·4) | 3 (2·4, 3·7) | 3·2 (2·1, 4·2) | 1440 |
| Non-Hispanic Black | 68 | 8 (6·2, 9·8) | 8·8 (6·6, 11) | 10·1 (5·8, 14·3) | 16742 |
| Hispanic and Black | 79 | 6·2 (4·8, 7·5) | 6.4(5 7.9) | 7·0 (4·7, 9·2) | 10616 |
| Integrated | 144 | 3·8 (3·2, 4·4) | 4.3(3.6 5) | 4·7 (3·4, 6·1) | 4395 |
| **Region**** | | | | | |
| Northeast | 97 | 8·3 (6·7, 9·9) | 8·4 (6·8, 10) | 9·4 (6·6, 12·3) | 9594 |
| South | 242 | 4·1 (3·6, 4·6) | 4·1 (3·6, 4·6) | 4·9 (4·1, 5·8) | 4786 |
| Midwest | 43 | 4 (2·8, 5·2) | 4 (2·8, 5·2) | 4·2 (2·3, 6) | 3891 |
| West | 137 | 3·8 (3·2, 4·5) | 3·9 (3·2, 4·5) | 4·1 (3·1, 5·2) | 4144 |
| **ZCTA poverty level #**** | | | | | |
| <10 | 104 | 3·4 (2·7, 4) | 3·6 (2·9, 4·4) | 3·5 (2·3, 4·7) | 1829 |
| 10 to <20% | 177 | 4 (3·4, 4·6) | 4·4 (3·7, 5·1) | 5·6 (4·3, 7) | 4390 |
| 20 to <30% | 132 | 4·9 (4·1, 5·7) | 5·3 (4·4, 6·3) | 5·9 (4·2, 7·5) | 6404 |
| ≥30% | 105 | 7 (5·7, 8·3) | 7·2 (5·8, 8·6) | 7·8 (5·5, 10) | 11691 |
| **ZCTA rural or urban status** | | | | | |
| Dense urban | 466 | 4·8 (4·4, 5·2) | 5·3 (4·8, 5·8) | 6·1 (5·2, 7·0) | 5123 |
| Metropolitan | 16 | 1·8 (0·9, 2·7) | 1·6 (0·8, 2·4) | 1·3 (0·4, 2·3) | 825 |
| Micropolitan | 19 | 3·2 (1·8, 4·6) | 2·9 (1·5, 4·3) | 2.5 (1·0, 4·0) | 1101 |
| Small town or rural | 17 | 3·2 (1·7, 4·7) | 2·4 (1·3, 3·6) | 2·1 (0·7, 3·6) | 991 |
| **Overall** | 519 | 4·4 (4·0, 4·8) | 4·7 (4·3, 5·2) | 5·4 (4·6, 6·2) | 5291 |

*different at p< 0·05 , ** different at p < 0·0001 for the sample population #Standardized to the U·S· dialysis population using all adults receiving dialysis for the treatment of end stage kidney disease on Jan 1st, 2017 identified through the United States Renal Data System database (N= 499 150)· &Standardized to the US population using ACS 2018 data ΩSeropositivity per 100 000 persons calculated as standardized count/category n X 100 000· +ZCTA Majority defined if population in ZCTA ≥ 60% Hispanic, Non-Hispanic Black, or Non-Hispanic white; if in remainder ZCTAs Hispanic and Black population exceeded ≥60%, ZCTA defined as 'Hispanic and Black', else as 'Other'·

**Supplemental Figure 1** Odds of SARS-CoV-2 seroprevalence by neighborhood and patient characteristics in January 2021, with comparison to July 2020

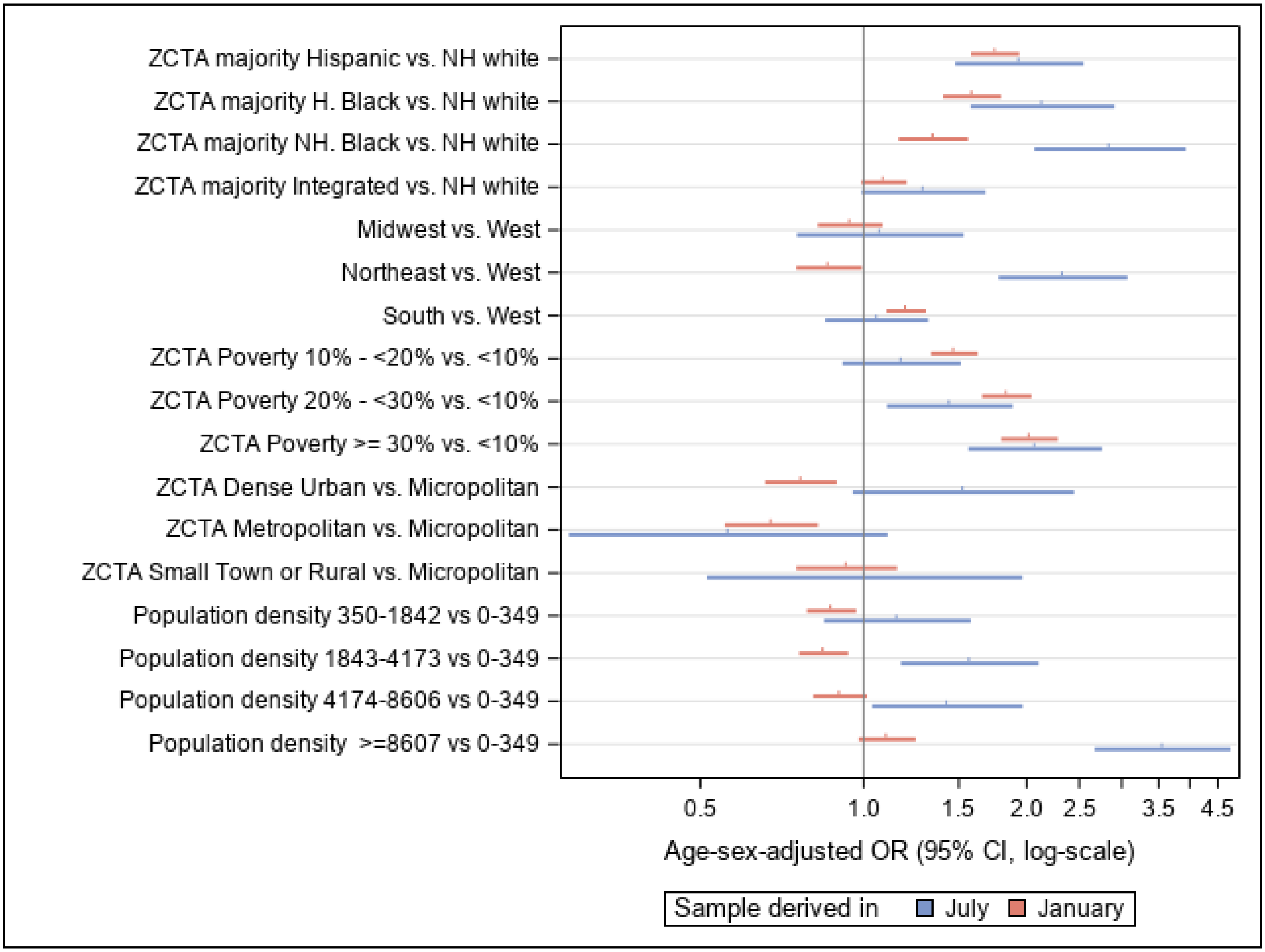


Compared with July data, the differences in seroprevalence by region, population density, and urban-rural status diminished. Persons living in majority Hispanic neighborhoods had higher odds of SARS-CoV-2 seroprevalence compared with persons living in majority white neighborhoods: OR 1.7 (95%CI 1.6, 1.9) in January 2021 versus OR 1.9 (95% CI: 1.5, 2.5) in July 2020. Persons living in majority Non Hispanic Black neighborhoods however had narrower differences in odds of seroprevalence compared with persons living in majority white neighborhoods: OR 1.3 (95% CI: 1.2, 1.6) in January 2021 versus OR 2.8 (95% CI 2.8, 3.9) in July 2020.

The differences by geographic region narrowed. The largest regional difference observed was 1.2 (95% CI: 1.1, 1.3) higher odds of seroprevalence in residents of the South versus West in January 2021, versus in July 2020 there was a 2.3 (1.8, 3.0) higher odds of seroprevalence in residents of the Northeast versus West. Persons living neighborhoods where ≥30% of residents had incomes below the federal poverty threshold still higher odds of SARS-CoV-2 seroprevalence: OR 2.0 (95% CI: 1.8, 2.3) in January 2021 versus OR 2.1 (95% CI: 1.6, 2.7) in July 2020.
